# Age-dependent risks of Incidence and Mortality of COVID-19 in Hubei Province and Other Parts of China

**DOI:** 10.1101/2020.02.25.20027672

**Authors:** Hongdou Li, Shuang Wang, Fan Zhong, Wuyin Bao, Yipeng Li, Lei Liu, Hongyan Wang, Yungang He

**Affiliations:** Shanghai Key Laboratory of Medical Epigenetics, International Co-laboratory of Medical Epigenetics and Metabolism Ministry of Science and Technology, Institutes of Biomedical Sciences, State Key Laboratory of Genetic Engineering and School of Life Science, Fudan University, Shanghai, China; Guanghan Research Center of Personalized Healthcare, Shanghai, China; Obstetrics Gynecology Hospital, The Institute of Reproduction and Developmental Biology, Fudan University, Shanghai, China

**Keywords:** Coronavirus, SARS-CoV-2, COVID-19, Incidence risk, Mortality risk

## Abstract

New coronavirus SARS-CoV-2 poses a big challenge for global public health in early 2020. Coronavirus Disease 2019 (COVID-19) caused by the virus rapidly spreads all over the world and takes thousands of lives in just two months. It is critical to refine the incidence and mortality risks of COVID-19 for the effective management of the general public and patients in the outbreak. In this report, we investigate the incidence and mortality risks of the infection by analyzing the age composition of 5319 infected patients, 76 fatal cases, and 1,144,648 individuals of the general public in China. Our result shows a relatively low incidence risk for young people but a very high mortality risk for seniors. Notably, mortality risk could be as high as 0.48 for people older than 80 years. Furthermore, our study suggests that a good medical service can effectively reduce the mortality rate of the viral infection to 1% or less.

## Introduction

On Jan 7, 2020, a new pathogenic virus caused pneumonia was identified in the sample of bronchoalveolar lavage fluid from a patient in Wuhan, Hubei province, China. It has typical features of the coronavirus family and therefore is classified as the subgenus Sarbecovirus, Orthocoronavirinae subfamily(1, 2). This virus has been named severe acute respiratory syndrome coronavirus 2 (SARS-CoV-2), and the disease it causes has been named “coronavirus disease 2019” (COVID-19). It is the third epidemic coronavirus that emerges in the human population in the 21st century, following the severe acute respiratory syndrome coronavirus (SARS) outbreak in 2002 and the Middle East respiratory syndrome coronavirus (MERS) outbreak in 2012(3, 4).

Coronavirus is one of the main pathogens of human respiratory infection owing to frequent cross-species infections. The emerging virus rapidly becomes a challenge for global public health due to spread by human-to-human transmission. The majority of the earliest COVID-19 patients were linked to the Huanan Seafood Wholesale Market. However, human-to-human transmission has frequently been occurring and that the epidemic has been gradually growing(5). As of March 4th, 2020, 80566 laboratory-confirmed cases have been reported in China. Internationally, more than 14396 cases have been reported in 77 countries(6, 7) The number of infected individuals is far surpassing that of SARS and MERS. SARS-CoV-2 can cause severe and even fatal respiratory diseases such as acute respiratory distress syndrome. It has been reported that SARS-CoV-2 is more likely to affect older males with comorbidities, suggests that age and comorbidity may be risk factors for poor outcomes(8, 9). In China, the reported death is approaching 3% in total of COVID-19 patients in the middle of February 2020.

At present, information regarding the prevalence and case-fatality on clinical features and epidemiology of COVID-19 remains scarce. However, a relatively accurate evaluation of incidence and mortality is required that will help refine the risk assessment and ensure that the public and patients are managed in an effective way. Therefore, it is necessary to quantitatively evaluate the risks for individual groups of different ages and gender. In this report, we show our initial analysis of the public data from local authorities. Our study shows that the incidence risk of COVID-19 might be as low as 0.1 for children while it could be over 0.9 for 40-years old adults. Our result also suggests that the mortality risk might be above 0.2 for patients older than 80 years old. Notably, the mortality risk is significantly different between patients of Hubei province and that of other parts of China.

## Method

### Data preparation

Basic information of COVID-19 cases was released on official websites by the national health commission of China and its local branches. A total of 5319 identified COVID-19 cases with 76 fatal cases were included in our analysis. All the 5319 COVID-19 cases were residents outside Hubei Province. Among the 76 fatal cases, 45 cases were reported as residents of Hubei province and 31 cases were reported as residents of other parts of China. Epidemiological characteristics, such as age, gender, and location were carefully checked to remove missing values or duplicated records. The composition of the age of the general public was obtained by data of 1,144,648 individuals from the General Census of China (2018)(10).

### Estimating incidence risk of COVID-19

We estimated the incidence risk of COVID-19 for different age groups in the general public by a maximum likelihood approach. In this approach, given the age composition of the general public and incidence risk of different age groups, age composition of COVID-19 cases can be obtained in

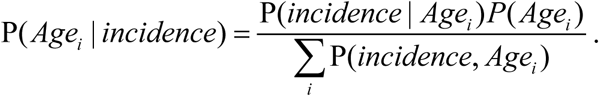

Where P(*incidence* | *Agei*) is the incidence risk of age group *i*; *P*(*Agei*) is the proportion of age group *i* in general public. We assumed that the incidence risk for different age groups could be obtained from a logistic function of age, 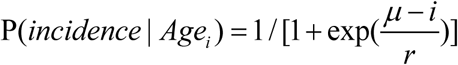. Likelihood of observation for age composition of 5319 COVID-19 cases can be maximized by searching for optimized μ and γ. Consequently, the incidence risk can be achieved in a maximum likelihood approach where the risk is given by a logistic function of age with estimated parameters μ and γ.

### Assessing mortality risk of COVID-19

To assess the mortality risk of COVID-19 in the general public, we used a maximum likelihood approach that is similar to that mentioned above. We obtained the age composition of fatal cases of COVID-19 as

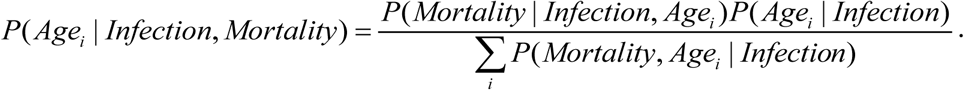

Where *P*(*Mortality* | *Infection, Agei*) is the risk of mortality condition on an individual’s age and infection state; *P*(*Agei* | *Infection, Mortality*) is age composition of fatal cases of COVID-19. In this study, we assumed that infection happens in all age groups for the general public and therefore have *P*(*Agei* | *Infection*) = *P*(*Agei*). We further applied the maximum likelihood approach to obtain the mortality risk of COVID-19 for different age groups in the general public *P*(*Mortality* | *Infection, Agei*). In the maximum likelihood approach, the mortality risk was given by the aforementioned logistic function of age but with mortality specific μ and γ. To eliminate the concern that the high mortality risk of older people may inflate the mortality rate of infected population, we further imputed the mortality rate of the infected people as 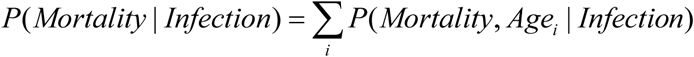.

## Result

### Characteristic of the COVID-19 cases and general public

The public data of a total of 5319 identified COVID-19 cases were included in our analysis. There were 2829 (53.2%) males and 2490 (46.8%) females in the COVID-19 cases, the male to female ratio turned out roughly equal across all age groups. The age of COVID-19 patients ranged from 0.5 to 97 years, with a mean of 45.2 years. The age and gender composition of COVID-19 patients and the public reference is presented in Table 1. Compared to the general public, the COVID-19 cases had higher average age, and there was a higher proportion of people aged 30 to 69 years.

**Table 1.** Age and gender composition of the general public and the identified COVID-19 cases.

| Age Groups | General public |  |  |  |  |  | COVID-19 cases |  |  |  |  |  |
| --- | --- | --- | --- | --- | --- | --- | --- | --- | --- | --- | --- | --- |
|  | Total | Male | Female | Total (%) | Male (%) | Female (%) | Total | Male | Female | Total (%) | Male (%) | Female (%) |
| 0-4 | 67393 | 35887 | 31506 | 5.89 | 3.14 | 2.75 | 51 | 24 | 27 | 0.96 | 0.45 | 0.51 |
| 5-9 | 63322 | 34279 | 29043 | 5.53 | 2.99 | 2.54 | 59 | 35 | 24 | 1.11 | 0.66 | 0.45 |
| 10-14 | 62248 | 33775 | 28473 | 5.44 | 2.95 | 2.49 | 55 | 32 | 23 | 1.03 | 0.60 | 0.43 |
| 15-19 | 58258 | 31552 | 26706 | 5.09 | 2.76 | 2.33 | 95 | 55 | 40 | 1.79 | 1.03 | 0.75 |
| 20-24 | 68050 | 36085 | 31965 | 5.95 | 3.15 | 2.79 | 239 | 140 | 99 | 4.49 | 2.63 | 1.86 |
| 25-29 | 92977 | 47710 | 45268 | 8.12 | 4.17 | 3.95 | 356 | 204 | 152 | 6.69 | 3.84 | 2.86 |
| 30-34 | 93201 | 46843 | 46358 | 8.14 | 4.09 | 4.05 | 524 | 291 | 233 | 9.85 | 5.47 | 4.38 |
| 35-39 | 81886 | 41517 | 40370 | 7.15 | 3.63 | 3.53 | 567 | 305 | 262 | 10.66 | 5.73 | 4.93 |
| 40-44 | 83574 | 42557 | 41017 | 7.30 | 3.72 | 3.58 | 579 | 349 | 230 | 10.89 | 6.56 | 4.32 |
| 45-49 | 102384 | 52108 | 50276 | 8.94 | 4.55 | 4.39 | 662 | 354 | 308 | 12.45 | 6.66 | 5.79 |
| 50-54 | 96850 | 48939 | 47911 | 8.46 | 4.28 | 4.19 | 631 | 319 | 312 | 11.86 | 6.00 | 5.87 |
| 55-59 | 69844 | 35208 | 34636 | 6.1 | 3.08 | 3.03 | 494 | 240 | 254 | 9.29 | 4.51 | 4.78 |
| 60-64 | 68014 | 34092 | 33923 | 5.94 | 2.98 | 2.96 | 349 | 157 | 192 | 6.56 | 2.95 | 3.61 |
| 65-69 | 54799 | 26974 | 27825 | 4.79 | 2.36 | 2.43 | 311 | 147 | 164 | 5.85 | 2.76 | 3.08 |
| 70-74 | 34810 | 16905 | 17905 | 3.04 | 1.48 | 1.56 | 153 | 85 | 68 | 2.88 | 1.60 | 1.28 |
| 75-79 | 22799 | 10745 | 12054 | 1.99 | 0.94 | 1.05 | 96 | 46 | 50 | 1.80 | 0.86 | 0.94 |
| 80-84 | 14845 | 6457 | 8389 | 1.3 | 0.56 | 0.73 | 57 | 25 | 32 | 1.07 | 0.47 | 0.60 |
| 85-89 | 6902 | 2870 | 4033 | 0.6 | 0.25 | 0.35 | 28 | 14 | 14 | 0.53 | 0.26 | 0.26 |
| 90-94 | 2031 | 665 | 1365 | 0.18 | 0.06 | 0.12 | 10 | 7 | 3 | 0.19 | 0.13 | 0.06 |
| 95+ | 458 | 131 | 327 | 0.04 | 0.01 | 0.03 | 3 | 0 | 3 | 0.06 | 0.00 | 0.06 |
| **Total** | **1144648** | **585299** | **559349** | **100** | **51.13** | **48.87** | **5319** | **2829** | **2490** | **100** | **53.19** | **46.81** |

We collected detailed information of 76 fatal cases and plotted the age composition of the cases and the general public in Figure 1. It is evident that death occurs more frequently in older people but rare for patients under 40 years old. The fatal cases were from 34 to 89 years old, with an average age of 71.47 and a standard deviation of 12.49.

**Figure 1.**
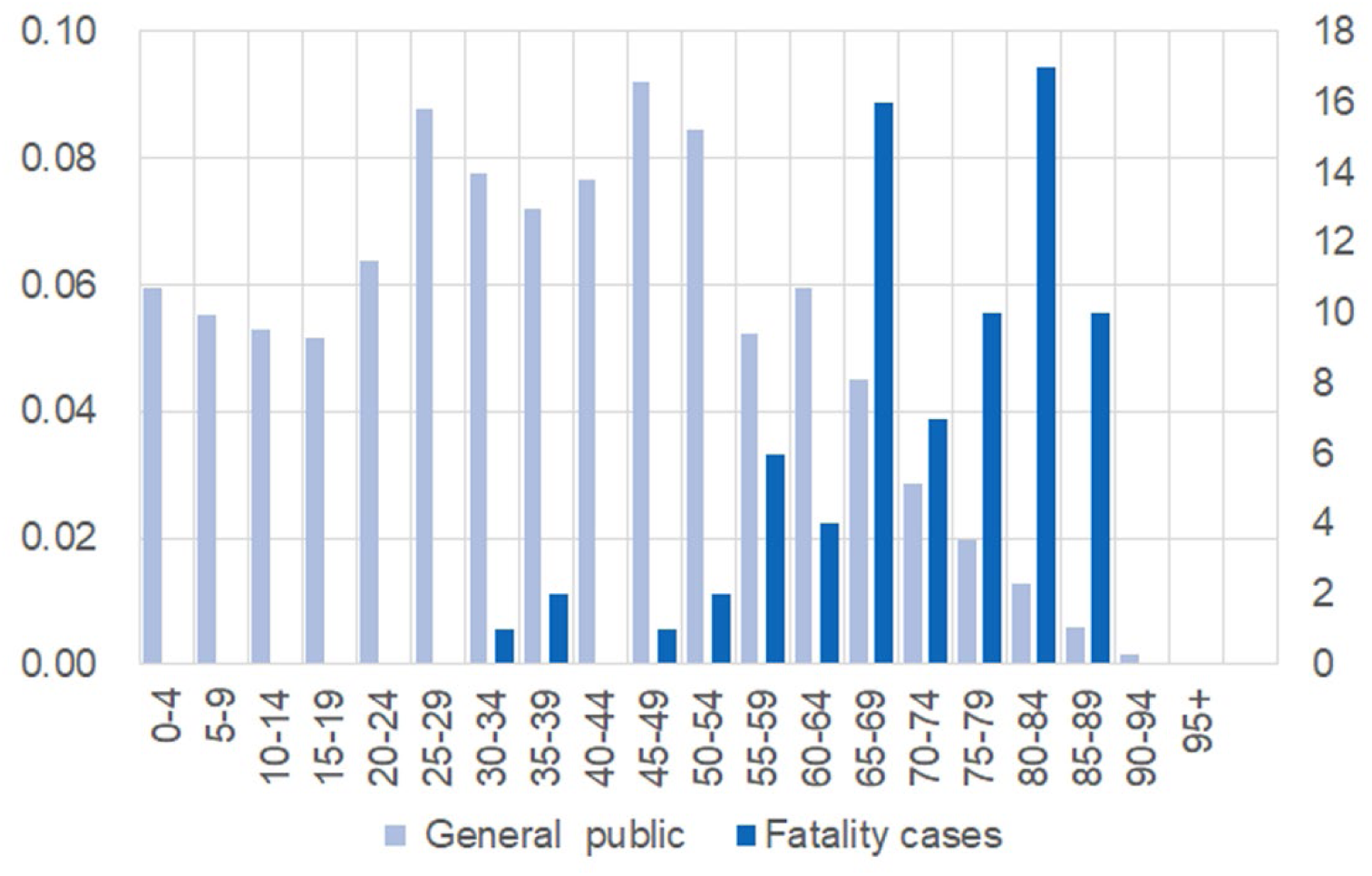
Different age composition between the general public and 76 fatal cases. Individuals were grouped and presented on x-axis. For the general public, proportion of each age group was shown on left-hand side; number of fatal cases in each group was present on right-hand side.

### Incidence risk of COVID-19 of the general public

Based on the age composition of 5319 COVID-19 cases and the 1,144,648 individuals of general public, we estimated the incidence risk in a maximum likelihood approach. Our result shows that the disease can happen in all age groups, and there lacks a significant difference between males and females (Figure 2). The difference in incidence risk for different gender is observed only for the groups between 15 and 50 years old. After the age of 15, males have a slightly higher incidence risk than women, but the increase is ignorable for people over 50 years old. Our result does not support a previous report that SARS-CoV-2 generally affects more males than females in the epidemic^8^. The incidence risk is low for children and teenagers but rapidly increases for adults. For adults over 40-years old, the risk is higher than 0.9 when they have full exposure to the virus.

**Figure 2.**
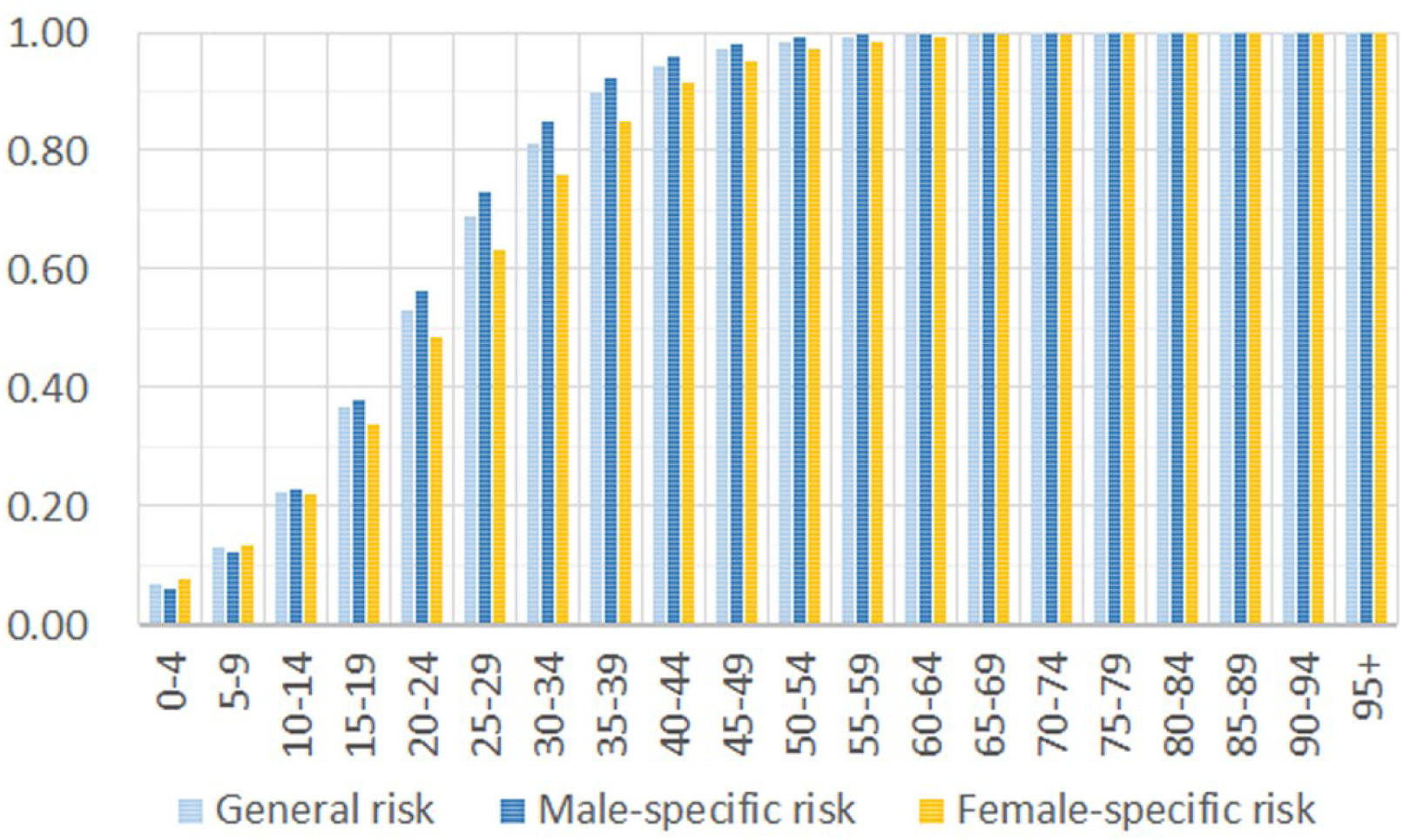
Incidence risk increases in older groups of the general public.

### Mortality risk of COVID-19 of general public

In our assessment for mortality risk, there is a significantly higher mortality risk in older adults (Figure 3). The estimated fatal probability is less than 0.01 for individuals under 40 years old, but it is more than 0.51 for people older than 90 years. The calculated risk is much higher than the previous reports. Our result is consistent with most of the earlier studies, supporting the hypothesis that older age is associated with an increased risk of mortality in COVID-19 patients. Our analysis of the total of 76 fatal cases suggests a mortality rate of 2.38% for general infection. However, we noticed that the mortality rate of COVID-19 in reports is significantly different between identified cases of Hubei province and that of other parts of China (11).

**Figure 3.**
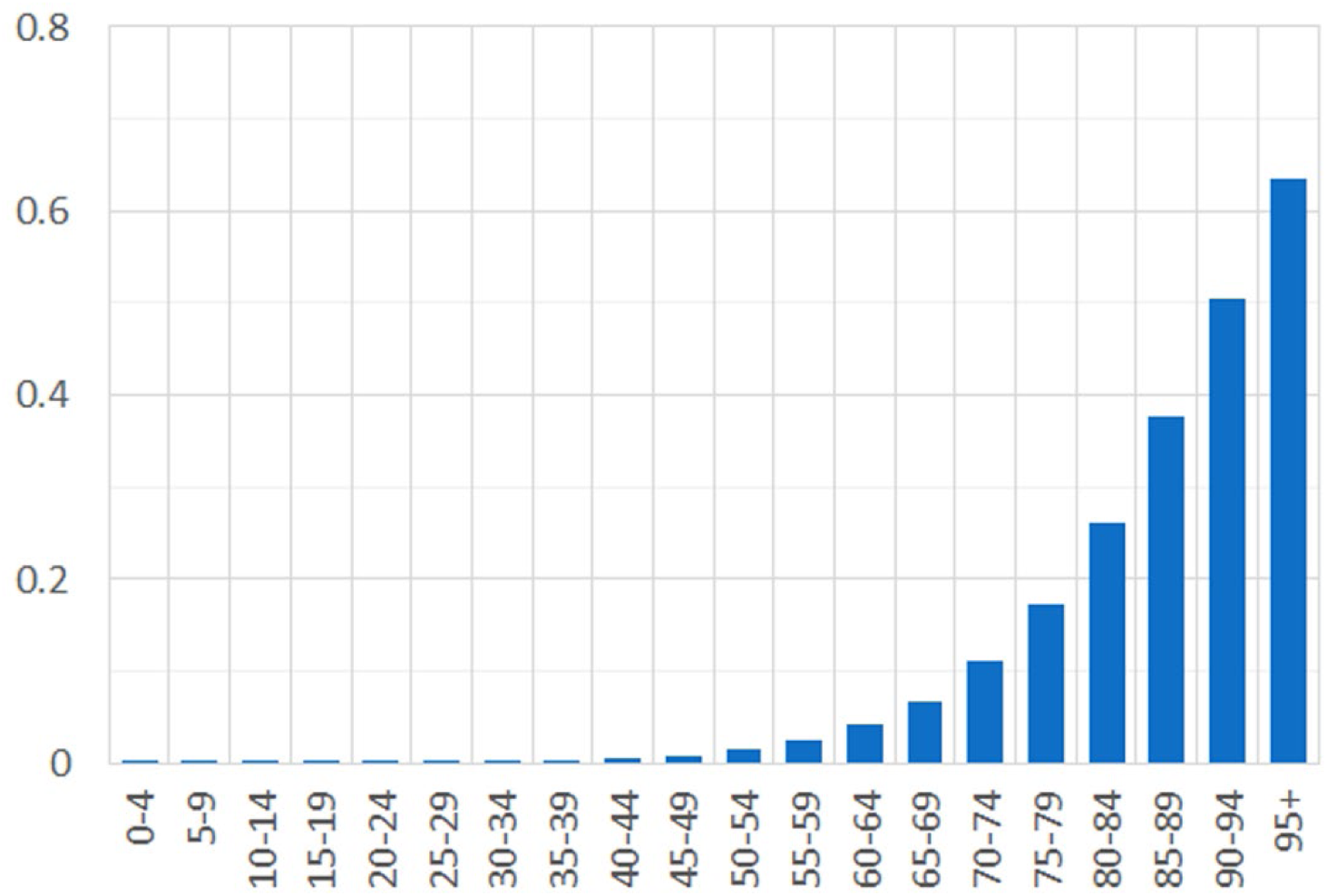
Mortality risk dramatically increases for older age groups.

### Different mortality risks in Hubei and other provinces

To compare mortality risk between Hubei and other provinces of China, we divided 76 fatal cases into two subsets, 45 cases from Hubei province and 31 cases from other parts of China. The aforementioned statistical analysis for mortality risk is applied to the two subsets with nine different age groups each. To account for variability, we further obtained a standard deviation of estimations by applying the same method on 1000 simulated data sets that generated with initial estimation. Our result shows that mortality risk is no more than 0.13 ± 0.10 for people over 80 years old outside Hubei province, but the risk is as high as 0.60±0.15 for the corresponding age group in Hubei province (Figure 4). Mortality risk falls under 0.05 for people younger than 70 years in other parts of China, while only people under 50 years old have a risk under 0.05 in Hubei province. We also calculated the expectation of mortality rate for a general infection inside and outside Hubei province as 4.78% and 0.95%.

**Figure 4.**
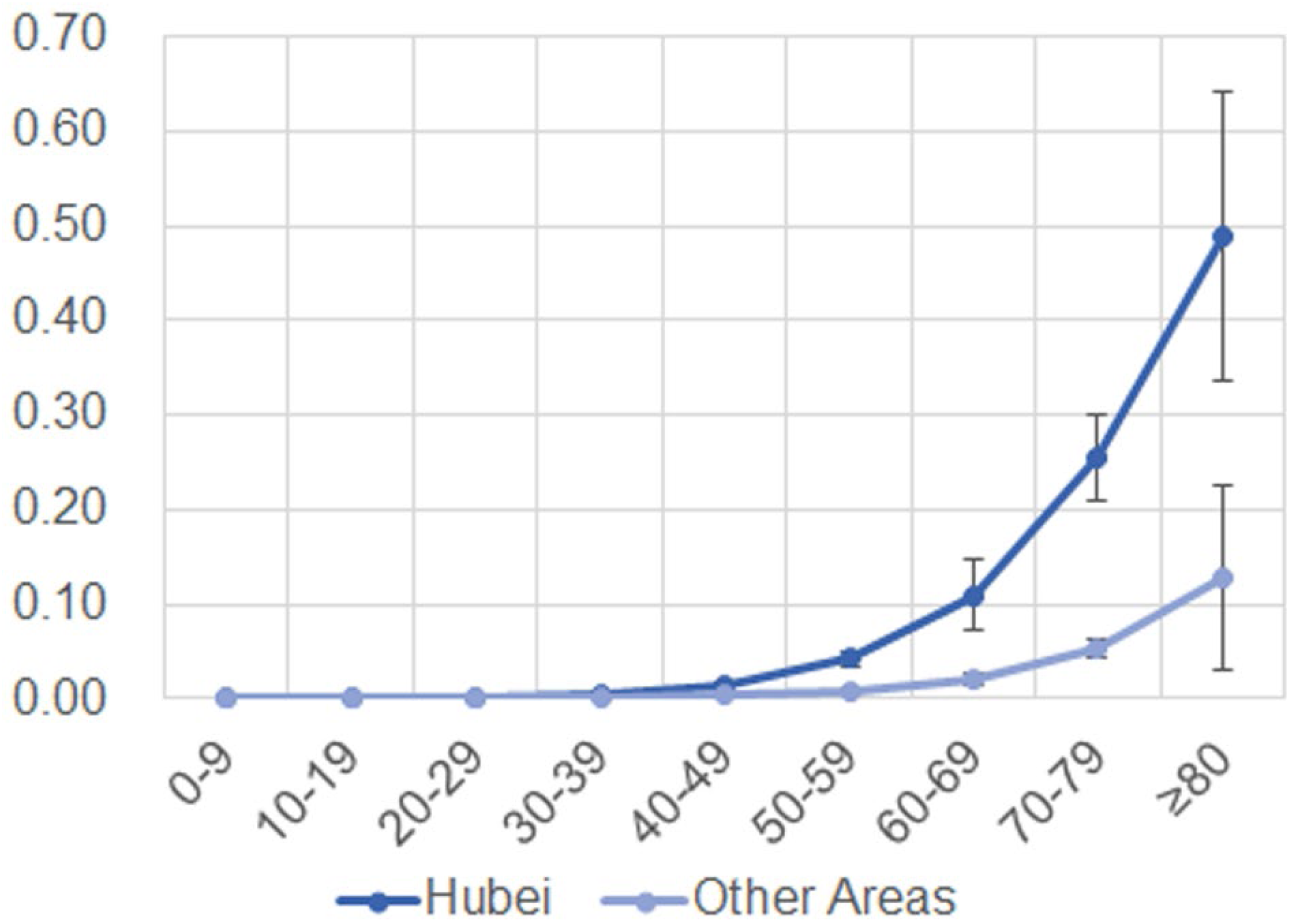
Estimated mortality risks in Hubei and other area of China. The risk was present on y-axis while age of grouped cases was shown on x-axis.

## Discussion

Our result suggests that there is a significantly higher mortality risk of COVID-19 for seniors than the previous report(11). It is probably due to the published report didn’t account for the increasing death of the identified COVID-19 cases. In the previous study, the crude mortality rate is only 2.3%, but the rate is approaching 3.7% on March 4th, 2020 (3,015 deaths among 80,566 identified COVID-19 cases). As of March 4th, 2020, there were still 5,952 COVID-19 patients in severe condition in China. It has been reported that the survival probability of critically ill patients continuously decreased with the increase of time since admission to the intensive care unit (12). Our analysis was based on the composition of the age of the different populations, and therefore it is less affected by the disease progression of patients, especially the increasing death of critically ill patients. Age has been reported as the independent predictor of adverse outcome in SARS and MERS. Comorbidities and low immune function of older people might be the major cause of higher mortality of coronaviruses(3, 4, 11). Prompt administration of antibiotics to prevent infection and strengthening of immune support treatment might reduce the mortality of seniors(8).

Our data showed that the mortality rate of COVID-19 is five times higher in Hubei province than that in other parts of China. On the one hand, this difference may be partially explained by insufficient medical resources due to such a large amount of patients in Hubei Province in the breakout. On the other hand, detailed information on the majority part of fatal cases (40 of 45 in total) from Hubei province was published before January 25^th^, 2019. The mortality rate of early reported cases may be overstated because case detection is highly biased towards the more severe cases. However, we strongly suggest international authorities try their best immediately to prevent too many COVID-19 patients overloading their health care system. Our hypothesis that a smooth-running health care system can effectively reduce the mortality rate of COVID-19 is strongly supported by the low mortality rate in other parts of China.

In conclusion, we investigate the incidence and mortality risks of the infection by analyzing the age composition of COVID-19 patients and the general public in China. Our data shows a relatively low incidence risk for young people but a very high mortality risk for old adults. Therefore, it is prudent to strengthen the tertiary preventive and clinical care of old aged patients to reduce mortality. Furthermore, our results also support that a good medical service can effectively reduce the mortality rate of the viral infection to 1% or less. Our study could be of value to medical authorities to implement effective medical service. The lack of complete data for all COVID-19 cases potentially increases the occurrence of selection and measurement biases in this study. Therefore, further large-scale epidemiological studies are necessary to elucidate the risk factors of COVID-19 for the general public.

## Data Availability

The data is avaliable upon request.

## Data Availability Statement

All datasets generated for this study are included in the article.

## Author Contributions

HY, HW, and LL contributed to the conception and design of the study. HL, SW, WB, and YL collected and analyzed data. HY, HL, and FZ wrote sections of the manuscript. All authors contributed to manuscript revision, read and approved the submitted version.

